# A feasibility study of a broadly applicable intervention to strengthen empowerment, self-management, and health among adults living with chronic illness in the United States

**DOI:** 10.64898/2026.07.07.26357498

**Authors:** Kisha N. Thompson, Marie Hamilton Larsen, Susan Hall, Dami Ko, Jørghild Jensen, Gyda Singstad, Kristin Heggdal

## Abstract

**Background:** Chronic illness is a major public health concern in Europe, the United States, and other high-income countries, limiting individuals’ capacity for self-management and health promotion. Empowerment interventions improve health outcomes while reducing healthcare utilization.

**Aim:** This study assessed the feasibility of implementing the Bodyknowledging Program, a broadly applicable health promotion intervention developed in Norway, at the community level in the US to evaluate participants’ experiences, program components, and self-management outcomes among adults living with chronic illness, and to identify the program’s strengths and areas for cultural adaptation to inform its cross-national transferability.

**Methods:** A multi-method feasibility design was used, including a group of participants living with various chronic illnesses. Reflexive thematic analysis was applied to analyze focus group data, examining participants’ experiences, program components, and outcomes. Facilitators’ field notes and post-intervention survey data were additional data sources.

**Results:** Three themes emerged through the thematic analysis: (1) acceptability of the BKP’s health promotion content and approaches among US participants, (2) implementation of the BKP intervention in a US community context, and (3) demand and ideas for continued implementation. Facilitator field notes identified challenges in implementing the hybrid format. Survey data confirmed that participants strongly agreed that the program enhanced their ability to recognize bodily signs and tolerance limits, manage symptoms, prevent deterioration, and promote their health. Participants reached consensus on the value of the program’s content, materials, organization, and communication strategies.

**Conclusion:** The Bodyknowledging Program is feasible and well-suited for implementation in the US. This community-based empowerment intervention leverages existing but unutilized human resources to strengthen self-management and health promotion among people with chronic illnesses across diagnostic categories. Further research across diverse settings is recommended to support broader dissemination.

## Introduction

The European Observatory on Health Systems and Policies [1] reports that more than one-third of Norwegian adults (37%) are diagnosed with at least one chronic condition, and 24–28% of adults report low quality of life due to health-related challenges. This situation is similar in other Nordic countries and has economic consequences, as the main share of total health budgets (70–80%) in this region is spent on treatment and care for people with chronic illness. Many countries in Europe and beyond share the same challenges due to the growing number of people diagnosed with chronic illnesses. In the US, 76% of adults report having one chronic condition, and 51% reported multiple chronic conditions [2]. Individuals with chronic illness face challenges in managing and adapting to physical, psychosocial, and life-course changes imposed by illness. These difficulties impact health perception, self-recognition, and well-being, often leading to uncertainty, social isolation, loss of control, and restricted life activities [3]. Strengthening individuals’ empowerment and self-management capacities is essential to promote health and ensure social functioning for this large number of people [4].

The World Health Organization (WHO) defines empowerment as both an individual and a community “process through which people gain greater control over decisions and actions affecting their health” [5, p.190]. The empowerment process has four key components: (1) patients’ participation through understanding their role, (2) patients’ knowledge acquisition to engage with their healthcare provider, (3) patient skills to partner in decision-making, and (4) a facilitating environment for open communication. Empowerment is crucial for individuals with chronic illness because it enables patients to take an active role in managing their health, equipping them with self-management skills, and promoting quality of life, thereby improving health outcomes and reducing healthcare utilization [6].

Empowerment interventions, including self-management support, are needed to address this public health challenge. Stepanian et al. [7] conducted a systematic review of 39 empowerment intervention studies and found that empowerment interventions critically improved self-management and health in chronic disease. Varela et al. [8] argue that empowerment interventions promote autonomy, competence, and relatedness, thereby renewing a sense of purpose and enhancing resilience and self-efficacy in persons living with chronic illness. Equipping patients with the tools to understand and manage the physical and psychosocial changes of chronic illness is fundamental to achieving these objectives [9]. The Bodyknowledging Program (BKP), developed in Norway [10, 11], is a broadly applicable intervention designed to support empowerment, self-management, health, and well-being in people living with chronic illness. The BKP is grounded in the premise that individuals possess internal health resources that can be strengthened both within the person and through healthcare interactions and used to promote well-being and health [12–14].

### Former research on the BKP in Norway

A study sample (n = 52) of adults aged 22 to 88 with various chronic illnesses (e.g., chronic obstructive pulmonary disease [COPD], heart disease, chronic inflammatory bowel disease [IBD], stroke, multiple sclerosis, or other neurological problems) participated in the first BKP trial [15]. The results captured participants’ experience of change in empowerment, self-management, and health. Participants described that “to be healthy within illness” implied coming out of a vacuum and participating in life again. This applied to various areas of life, including family, work, and social life, and showed that the BKP pays particular attention to psychosocial dimensions. Subsequently, the BKP was tested in a community-based sample (n = 11) who were aged 30 to 60. Results aligned with the first trial and additionally showed that the intervention contributed to participants’ regaining control of their life situation [16]. A third study (n = 108, aged 21–89) compared patient outcomes in specialist and community care by measuring changes in individual sense of coherence during the BKP intervention [13]. The results showed a combined clinically significant increase in sense of coherence scores, with a larger mean change in the community care group. A fourth study [17] tested the efficacy of BKP in a community-based sample (n = 37), with an average age of 54, using the Outcome Rating Scale and found significant changes in recovery and health throughout the program period. The greatest change was in the personal and general well-being dimensions, with dysfunctional patients moving from below to above the ORS cut point of 25, indicating better coping, well-being, and recovery.

Two qualitative studies [18, 19] were conducted to explore the feasibility of the BKP from the perspectives of healthcare professionals in Norway. Both studies showed that the BKP assisted professionals in addressing the whole person and individual needs and facilitated empowerment and patient activation. The professionals described being challenged to develop new competencies that required a major change in their professional role to promote participant activation and facilitate individuals’ health-promotion processes. Findings from all these studies provide evidence that the BKP is broadly applicable to strengthening empowerment, self-management, and health among individuals with chronic illness across diagnostic categories, ages, genders, and clinical sites. While the BKP has proven effective in Norway, its feasibility in other cultural contexts remains unexplored. The aim of this study was to assess the feasibility of implementing the BKP at the community level in the US; evaluate participants’ experiences, program components, and self-management outcomes among adults living with chronic illness; and identify the program’s strengths and areas for cultural adaptation to inform its cross-national transferability.

## Materials and methods

### Design overview

This multi-method feasibility study evaluated a 7-week pilot of the BKP in the US using a one-group pre-post intervention design, facilitated by two registered nurses trained in leading the BKP. Three primary data sources informed the evaluation: (1) a post-intervention focus group with participants, (2) facilitator field notes documented after each weekly session, and (3) a brief mixed quantitative and qualitative post-intervention survey. Data were collected and analyzed independently, then integrated to enhance credibility and provide contextualized findings from multiple perspectives [20]. The qualitative analysis generated an in-depth understanding of participants’ and facilitators’ experiences, which was compared with survey findings to inform feasibility conclusions and identify pragmatic adaptations for implementing the BKP in the new cultural context [21]. Quantitative analyses of validated instruments and program outcome data are reported separately.

### Participant recruitment and sampling

The 7-week intervention took place from September to October 2024, with a convenience sample of community-based adults in New York City suburbs. Community partnerships and flyers were used to recruit participants. Interested parties underwent telephone screening for enrollment. Eligible participants were over 18 years old, had a chronic illness, sought self-management support, and were able to participate in group discussions. Individuals were excluded if they were receiving palliative cancer care, were experiencing an acute medical crisis, or had attended support groups within 3 months prior to enrollment. Twenty participants were screened; 18 were eligible and participated in the BKP; 16 completed the intervention (attending at least 5 of 7 sessions) and a post-intervention survey. Nine participated in the focus group.

### Intervention: The Bodyknowledging Program

The first three group sessions were held in person; the last four were conducted online. The program approach includes structured dialogue, group work, facilitated discussions, physical activity, and reflective activities such as journaling (Table 1). The BKP aims to leverage participant expertise to promote chronic illness self-management by applying salutogenic [22] and person-centered principles [23]. Participants’ inherent knowledge of illness and health is acknowledged and utilized as a key resource (i.e., awareness of personal tolerance limits for activity; physical and psychosocial factors; social interactions on uncovering factors that make the illness better or worse, and possibilities to be active and participate in society) [11, 14].

**Table 1.** Structure, Content, Pedagogical Methods, and Tools of the Bodyknowledging Program.

|  |  |
| --- | --- |
| **Session 1:** Program Introduction | Information about program structure, content, and pedagogical tools. Short introduction to the Bodyknowledging Model. |
| **Session 2:** Uncertainty | Participants' own themes about living with chronic illness, especially the phase of uncertainty—escaping and denying the sick body.<br>Introduction to physical exercises and the use of the booklet/diary. |
| **Session 3:** Loss of Life Space | Introduction to the phase of Losing life space—grieving and anger.<br>Participants' own themes concerning challenges with a long-term condition and their health promotion strategies.<br>The use of booklet/diary between and in sessions.<br>Physical exercises. |
| **Session 4:** Loss of Life Space (in-depth work) | In-depth work on the phase of Listening and understanding the body's signs—strengthening hope.<br>Acknowledging their body as a source of knowledge about health.<br>Exploring their body's limits of tolerances concerning the type and magnitude of physical activity (i.e., work, workout), essential factors of the physical environment with an impact on their health and type and magnitude of social relations (social network, significant others, work-relations).<br>Communicating their limits of tolerance to others. Participants' own themes and strategies concerning health promotion in chronic illness.<br>The use of booklet/diary.<br>Physical exercises. |
| **Session 5:** Listening and Understanding the Body's Signs (introduction) | Participants' own themes and strategies for creating health within illness.<br>How to get to know one's limits of tolerance for activity, factors of the physical and social environment.<br>Encounters with others. |
|  | Exploring the possibilities for extending their limits of tolerances in order to facilitate wellness.<br>The use of booklet/diary.<br>Physical exercises. |
| **Session 6:** Listening and Understanding the Body's Signs (in-depth work) | In-depth work on getting to know one's body's capacity.<br>Introduction of the phase of Integrating embodied knowledge—exploring new possibilities for health.<br>Living with chronic illness and encountering significant others, work relations and the society.<br>Participants' own themes of health promotion in chronic illness.<br>The use of booklet/diary.<br>Physical exercises. |
| **Session 7:** Integrating Knowledge – New Possibilities for Well-being and Health | Summary of the Bodyknowledging Model in relation to each participant.<br>Encounters with society.<br>Participants' thoughts and actions on how to sustain and strengthen their health after program completion.<br>Physical exercises. |

### Ethical Considerations

Ethical approval was obtained from Northeastern University in Boston, MA, USA (IRB #24-02-11). All participants provided written informed consent to participate in the research after receiving comprehensive information about the study. To ensure confidentiality, personal identifiers were removed from the data before analysis. Electronic data were stored on password-protected, university-approved platforms, while physical documents were kept in a locked cabinet separate from study data and destroyed after study completion. A small financial incentive was provided for intervention participation and survey completion; however, no additional incentive was offered for participation in the focus group.

### Data Collection

At 1-week post-intervention, the optional 90-minute online focus group was conducted using a semi-structured interview guide (S1 File). The guide used open-ended questions to explore the program’s structure, content, participants’ experiences, acceptability, and areas for improvement. Prompts were used to elicit more context-rich responses. The session was recorded and transcribed via Zoom for Teaching and Learning (version 6.0). The transcript was verified, de-identified, and distributed to the team for analysis. After each weekly session, facilitators recorded reflective field notes that captured group dynamics, facilitation insights, and program considerations. During Session 7, a post-intervention survey was administered via Qualtrics (Provo, UT); it included 36 items covering demographics, program evaluation, satisfaction, participation patterns, and feedback, as well as 7 open-ended questions.

### Data Analysis

Qualitative analysis of the focus group transcript was adapted from Braun and Clarke’s [24] reflexive thematic analysis approach, a method for identifying recurring patterns, insights, and experiences (Table 2), using NVivo [25]. This process employed a hybrid inductive-deductive approach, with inductive themes reflecting participants’ perspectives, which were further analyzed using Bowen et al.’s [26] feasibility domains: acceptability, demand, implementation, practicality, and adaptation. Domains such as fidelity, integration, and limited efficacy testing were excluded as being beyond this evaluation’s scope. Participants’ experiences of the BKP were prioritized in presenting the findings. Furthermore, the facilitators’ field notes were sorted under relevant feasibility themes. The quantitative data from the post-intervention survey were analyzed in IBM SPSS Statistics (version 28) using descriptive statistics, and the seven open-ended questions were also analyzed thematically.

**Table 2.**
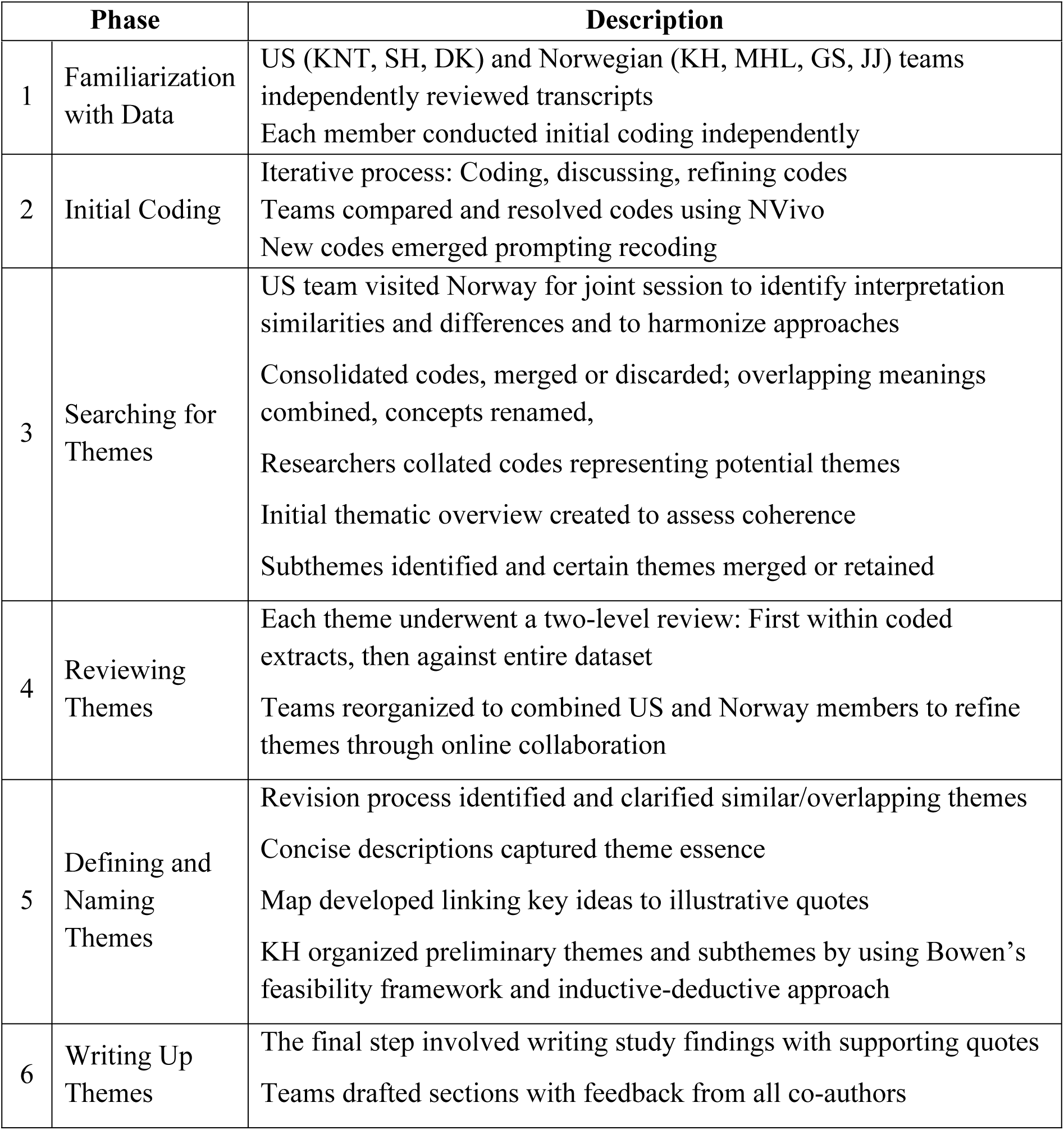
Overview of the Focus Group Transcript Reflexive Thematic Analysis Process.

| Phase | Description |
| --- | --- |

|  |  |  |
| --- | --- | --- |
| 1 | Familiarization with Data | US (KNT, SH, DK) and Norwegian (KH, MHL, GS, JJ) teams independently reviewed transcripts<br>Each member conducted initial coding independently |
| 2 | Initial Coding | Iterative process: Coding, discussing, refining codes<br>Teams compared and resolved codes using NVivo<br>New codes emerged prompting recoding |
| 3 | Searching for Themes | US team visited Norway for joint session to identify interpretation similarities and differences and to harmonize approaches<br>Consolidated codes, merged or discarded; overlapping meanings combined, concepts renamed,<br>Researchers collated codes representing potential themes<br>Initial thematic overview created to assess coherence<br>Subthemes identified and certain themes merged or retained |
| 4 | Reviewing Themes | Each theme underwent a two-level review: First within coded extracts, then against entire dataset<br>Teams reorganized to combined US and Norway members to refine themes through online collaboration |
| 5 | Defining and Naming Themes | Revision process identified and clarified similar/overlapping themes<br>Concise descriptions captured theme essence<br>Map developed linking key ideas to illustrative quotes<br>KH organized preliminary themes and subthemes by using Bowen's feasibility framework and inductive-deductive approach |
| 6 | Writing Up Themes | The final step involved writing study findings with supporting quotes<br>Teams drafted sections with feedback from all co-authors |

### Validity, Reliability, and Methodological Integrity

The semi-structured interview guide and post-intervention survey were standardized BKP tools previously piloted and shown to be reliable, then adapted for clarity and cultural appropriateness. Analytical trustworthiness and credibility were strengthened through multiple validation strategies, including an audit trail documenting coding decisions, dual independent coding with consensus resolution, and reflexive journaling to address researcher bias [27]. The researchers’ personal and professional backgrounds influenced the study’s approach, facilitating community engagement and recruitment as well as providing theoretical and practical expertise in chronic illness self-management. The research team carefully considered transferability across cultural and organizational contexts, maintaining a reflexive awareness that participants possess experiential health knowledge that is often underutilized in traditional healthcare. This reflexive stance focused on the program’s feasibility toward health empowerment processes.

## Results

The BKP final sample of 16 participants had an average age of 65 (SD = 14.5), with 68% (*n* = 11) retired, 50% (*n* = 8) identifying as Black, and one male participant (Table 3). Participant health concerns included asthma, diabetes, high blood pressure, kidney disease, autoimmune illnesses, mobility limitations, chronic pain, obesity, depression, and memory loss. Half (*n* = 8) reported completing education at the bachelor’s degree or higher level. The sample demographics showed varying impacts from social and political determinants of health. Several participants reported needing outside assistance to meet basic financial needs, experiencing housing insecurity, or lacking healthcare coverage. Some also noted the limited availability of support from others. Those who had support systems identified children, friends, siblings, a pastor, and a girlfriend as key sources of support.

**Table 3.**
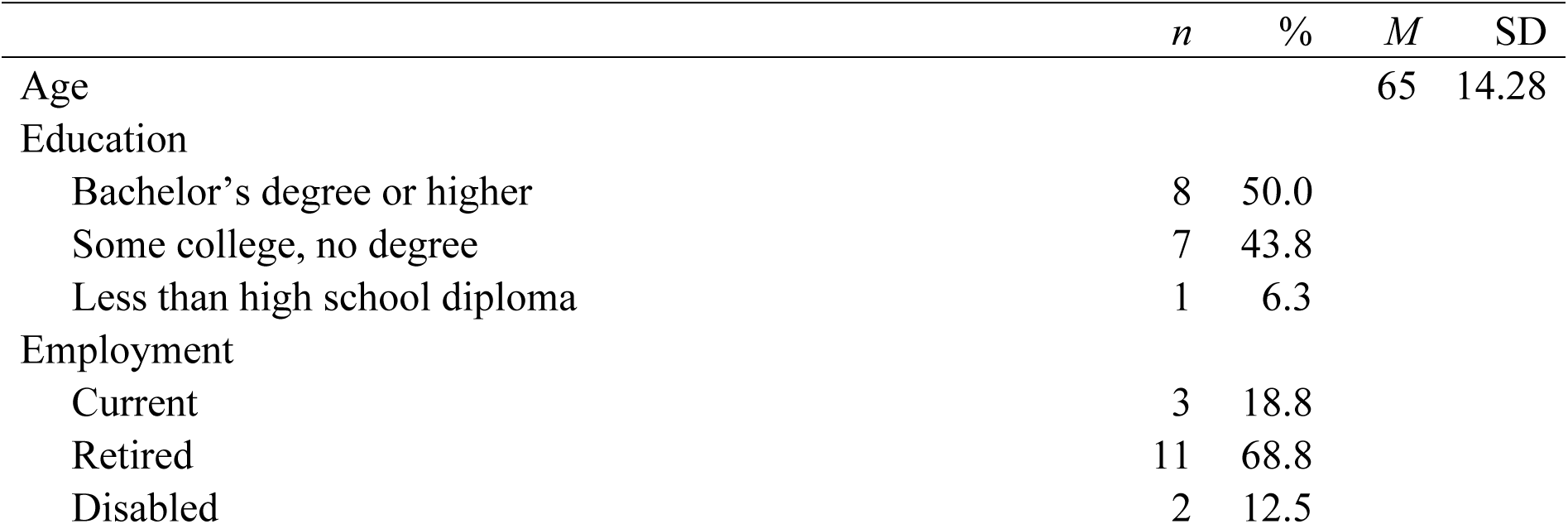
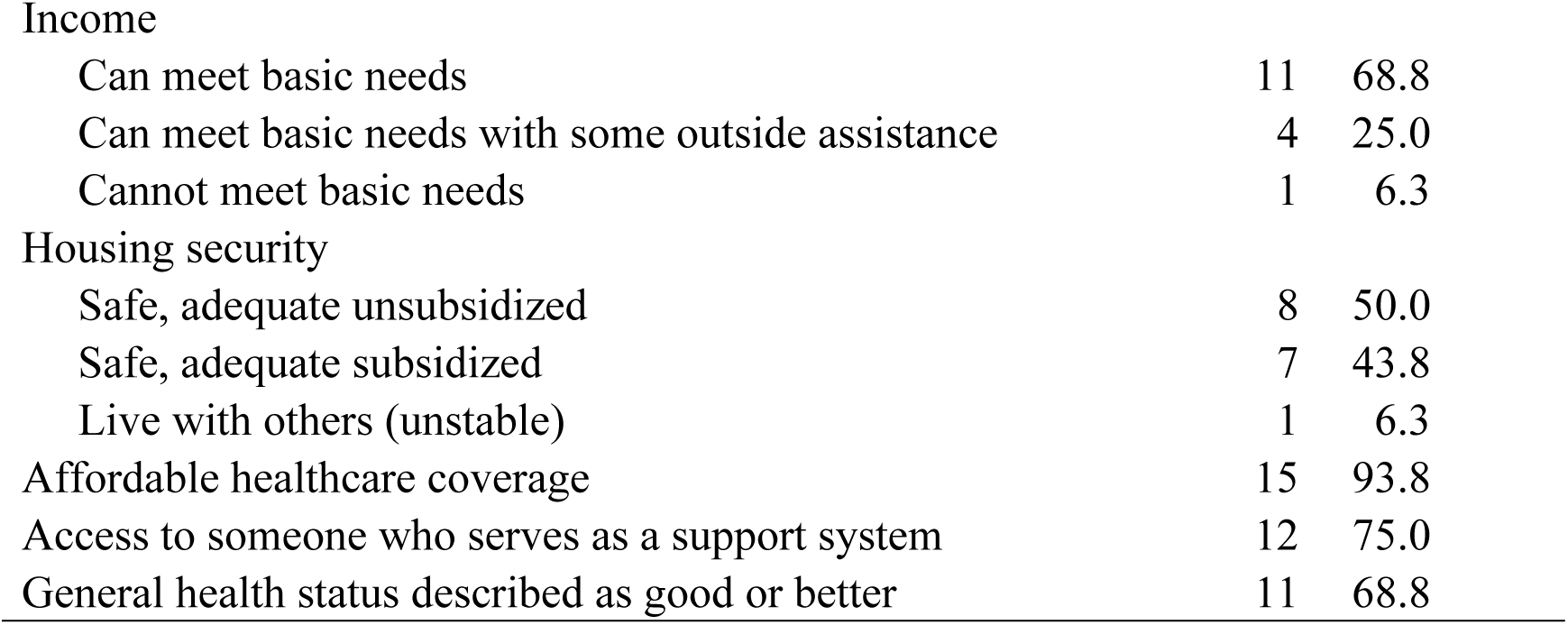
Post-intervention Evaluation Survey Participant Characteristics (N = 16)

|  | *n* | % | *M* | *SD* |
| --- | --- | --- | --- | --- |
| Age |  |  | 65 | 14.28 |
| Education |  |  |  |  |
| Bachelor's degree or higher | 8 | 50.0 |  |  |
| Some college, no degree | 7 | 43.8 |  |  |
| Less than high school diploma | 1 | 6.3 |  |  |
| Employment |  |  |  |  |
| Current | 3 | 18.8 |  |  |
| Retired | 11 | 68.8 |  |  |
| Disabled | 2 | 12.5 |  |  |

|  |  |  |
| --- | --- | --- |
| Income |  |  |
| Can meet basic needs | 11 | 68.8 |
| Can meet basic needs with some outside assistance | 4 | 25.0 |
| Cannot meet basic needs | 1 | 6.3 |
| Housing security |  |  |
| Safe, adequate unsubsidized | 8 | 50.0 |
| Safe, adequate subsidized | 7 | 43.8 |
| Live with others (unstable) | 1 | 6.3 |
| Affordable healthcare coverage | 15 | 93.8 |
| Access to someone who serves as a support system | 12 | 75.0 |
| General health status described as good or better | 11 | 68.8 |

The participant focus group and HCP field notes offered valuable insight into the intervention’s feasibility, acceptability, and practical implementation. The combined data sources provided a nuanced understanding of how the intervention was received and which areas needed improvement. Three main themes captured the feasibility of the intervention: (1) Acceptability of the BKP’s health promotion content and approaches among US participants; (2) Implementation of the BKP intervention in a US community context; and (3) Demand and ideas for continued implementation, containing a total of 11 subthemes (Table 4).

**Table 4.** Overview of the Main Themes and Subthemes.

| **Deductive Main Themes** | **Inductive Subthemes** |
| --- | --- |
| Acceptability of the BKP's health promotion content and approaches among US participants | The message of self-empowerment is a valuable component<br>Learning tools are catalysts for self-reflection and empowerment<br>Discovering one's own health promotion process involves acknowledging one's own body and skill application<br>The strength of social interaction and support increases when sharing and learning together with peers |
| Implementation of the BKP intervention in a US community context | The program was well prepared and planned<br>The facilitator's compassionate presence is essential to participant experience<br>Physical and psychological barriers for implementation require creative solutions |
|  | The online format has advantages and challenges |
| Demand and ideas for continued implementation | Incentives impacted participation.<br>Participants expressed a need for continuation and ideas for recruitment.<br>Suggestions for program change, additional content, and methods were offered |

### Theme 1: Acceptability of the BKP’s health promotion content and approaches among US participants

This theme explores how the participants and facilitators involved in the BKP pilot assessed the content and pedagogical approaches as suitable, satisfying, or attractive, and how the participants assessed the outcomes. Four subthemes serve to elaborate on the main theme: **The message of self-empowerment is a valuable component.**

Participants highlighted self-empowerment as a central and valuable aspect of the BKP, involving learning to listen, understand, and utilize their inherent, experience-based knowledge of health and illness as a tool for health promotion. This was highlighted as a new experience in the encounter with healthcare:

> We don’t get that message of self-empowerment strongly enough when we are in the regular sort of medical system. There’s very much emphasis on medication and procedures of more specialists, just this, while simple messages of taking things into your own hands as far as just listening to your own body and making your own choices based on paying attention is left out.

The rainbow model (S3 Fig) in the poster and booklet was described as a means for participants to self-assess their current status and maintain wellness, allowing them to identify their position in their health promotion process and track their progress. Facilitators noted how readily the US participants engaged with the model, often sharing their self-assessments from the first session onward and encouraging others to do the same. One of the facilitators reflected that even participants who were initially hesitant to join eventually opened up, sharing in depth experiences of illness, loss, and grief—particularly during Sessions 3 and 4, which focused on the “blue phase” (i.e., losing life space, grieving, and anger).

#### Learning tools are catalysts for self-reflection and empowerment

Participants assessed the educational tools, particularly the booklet and the rainbow model poster, as attractive and helpful in facilitating their own health promotion process. The booklet’s diary format and reflective questions encouraged participants to track progress, recognize change, and make sense of their experiences living with chronic illness. One participant described it as “a personal diary” that reinforced learning between sessions and served as a lasting record of growth. Another participant shared:

> I thought the questions were very good because it made you self-reflect, to see where I started and where I ended. I could see progression, or how I looked at my Bodyknowledging differently, and how I were able to have a strategy now to deal with what I’m going through, not as opposed before. When I look at my notes, it helped me realize that, okay, I’m moving forward, I get to another phase.

Participants conveyed that the learning activities served as a means of recognizing challenges in living with chronic disease and gaining new insights into their own health processes. The exercises encouraged reflection, mutual support, and the exchange of coping strategies among peers. By applying BKP principles, participants reported becoming more aware of their physical and emotional limits and learning to act on them in everyday life.

Structured learning activities helped participants articulate what concepts such as uncertainty, loss, or health meant to them and stimulated fruitful group discussions and shared learning. The relaxation exercises promoted presence and balance, although participants’ experiences varied. Some needed time to feel confident using the technique, as one participant shared:

> The first day I tried to do it, I could not concentrate, but after two, three days, I said to myself: I better concentrate, keep doing it, and see if it’s work. So, I tried to do it at home, and it worked. It’s helped me. I turn off the TV when I’m doing it, and I even pull down the shade. And then I stay in my room. I do my little prayer, and then I start doing it. That’s all I have to do. Stay in a quiet place, calm, quiet, and then it will work. I don’t know if it’s some kind of magic thing, but I find out that the pain on my knees is gone, and I am able to walk around more now, so I think that relaxation worked for me.

The group’s willingness to share was a pleasant surprise to the HCPs; likewise, the interaction among participants was comforting and supportive. The “Enough is enough” learning activity was highlighted as being particularly powerful, allowing participants to explore personal boundaries and communicate them openly. Facilitators also noted strong engagement among BKP participants in this activity, even though some voiced that the online format limited sharing of experiences.

The pilot started with 18 and ended with 16 participants, although attendance varied slightly between sessions. Participants viewed the group size as manageable but suggested that smaller breakout groups could enhance discussions and interaction. Differences in attitudes towards technology emerged. Some participants felt that limited digital literacy among older adults could hinder participation in online sessions, while others argued that motivation, not age, determined one’s ability to use technology. As one participant illustrated:

> It has nothing to do with age. I’m going to be [age] years old at the end of this year in December and I have been doing Zoom and other technologies for years. So, I say, don’t put your age in the way of it. It has nothing to do with your age. It has to do with your needs and your desires.

Overall, motivation and support for technology use were emphasized as prerequisites for successful future implementation. This is further elaborated in the presentation of the third theme.

#### Discovering one’s own health promotion process involves acknowledging one’s own body and skill application

Participants emphasized the BKP was transformative in developing individualized approaches to health promotion by acknowledging their bodies and applying self-care skills. The program helped them interpret physical responses and recognize how interpersonal relationships directly impacted their symptoms and overall well-being. As one participant explained:

> I think that the rainbow model identifies the phases that you go through and the strategies you could use to get through a stage…so that you don’t have to stay stuck because now you can identify what’s really happening with you.

This heightened somatic awareness encouraged participants to set boundaries, disengage from draining relationships, and prioritize well-being, all of which were experienced as a shift toward self-advocacy and personal empowerment. Many integrated new practices such as walking, meditation, and outdoor activities into their daily routines, reporting improved physical and mental health. One participant enthusiastically shared the progress:

> Before, I could not walk one block. I had to sit down for a few minutes and take my balance back, and then do it again. And you will not believe it. On Sunday, I walked from my house, all the way to the mall. I went alone and I didn’t feel any pain. I don’t know if it’s relaxation. I don’t know what it is, but I did it.

Facilitators observed participants’ growing confidence as they took practical steps toward personal goals, from pursuing creative projects to planning community initiatives. These reflections illustrated the widening of the participants’ life space and their emerging ability to act on newfound insights, thus signaling meaningful changes in self-agency and health behavior.

#### The strength of social interaction and support increases when sharing and learning together with peers

Participants described social interaction and mutual support as key benefits of the BKP. Sharing experiences with peers facing similar challenges created a sense of belonging, reduced feelings of isolation, and fostered emotional relief. Being listened to and understood was emphasized as deeply validating and “humanizing” because it offered a safe space to discuss issues they seldom shared elsewhere. As one participant revealed:

> It’s the really deep stuff of human experience, the most painful, you know, that really makes life poignant, and to be able to share it in a group setting is so validating and just humanizing because I think maybe some of us feel very alone in our illness. Because in general society, you know, we don’t usually talk about those things a lot. And it’s not socially acceptable to talk about it in happy gatherings when you get together with friends. This group provided the experience of sharing, you know, it slowed everything down, and then we got to talk with each other and provide support to each other.

Mutual peer support and facilitator support were described as having a healing effect, and participants expressed a desire to continue the group to have more of that experience. Unlike traditional support groups, the BKP environment emphasized personal strengths and wholeness rather than illness. Participants valued giving and receiving support and described learning from each other’s experiences as empowering. Witnessing their peers’ courage and coping strategies helped them rediscover their own inner resources, fostering a sense of meaning, connection, and resilience. This was an opportunity for participants to discover and learn from their own and their peers’ experience-based knowledge. One participant put it this way: “There’s this inner knowledge, the survival knowledge, and I feel my peers’ strength, and it’s given me strength.”

### Theme 2: Implementation of the BKP intervention in a US community context

Participants and facilitators highlighted key factors for the successful delivery of the BKP intervention in a community setting, such as a US public library; reported on the format’s functionality and implementation challenges; and offered recommendations for future implementation. The theme included four subthemes: (1) Program well prepared and planned; (2) Facilitators’ crucial role, presence, and compassion; (3) Advantages and challenges of the online format; and (4) Physical and psychological barriers for implementation requiring creative solutions.

#### The program was well prepared and planned

Participants assessed the implementation of the BKP as successful and attributed this success to the program’s preparation, format, and materials as well as the facilitators’ compassion and approaches. The seven-week structure—with three in-person sessions followed by four online sessions—was considered both feasible and effective. Meeting in person at the start was seen as important for building trust and comfort, enabling deeper sharing in later online sessions. While participants found the online sessions convenient, they emphasized the need for technical guidance to ensure equal participation. As one noted, “It is important to let people know that you’re gonna provide an opportunity to get to know online and to become more comfortable with it at whatever level they’re at.” The group process was highlighted as engaging and supportive, fostering openness and reflection. One participant noted;

> “By the seventh week, we were intertwined.” The group process helped them share their experiences and learn how to handle their health-related challenges. The facilitators could then facilitate the learning process by confirming the participants’ experience, offering support, and challenging them to explore more of the health promotion factors in their lives and how they could act in the direction of health and well-being: Something happened during the sixth session: Some participants started to share their dreams for the future. The dialogue on this was exciting. I challenged these participants to think about how they could take a small step towards their dream and share something about this in the seventh session”.

#### The facilitator’s compassionate presence is essential to participant experience

Participants highlighted the facilitators’ presence and compassion as essential to the BKP, and one participant argued that “compassion carries the program.” They described the facilitators as fully engaged, careful with their words, and skilled at creating an inclusive atmosphere where everyone felt welcomed and was given “a voice.” One of the participants expressed it this way:

> You had such a good way of acknowledging everyone in the class. You let us speak out of turn. The diplomacy that you exhibited was something that I think most people didn’t realize that you were doing. But you just made everybody feel that they were correct, that everyone was right. Everyone felt welcome. You didn’t feel like you’re being singled out for anything unless it was positive. And that’s huge. That’s a talent.

Participants emphasized the facilitators’ kindness, lack of superiority, and focus on empowerment and mutual support, which helped the participants share difficult stories and feel safe in the room.

Many wanted the group to continue after the program and stressed that its effectiveness depended on having trained facilitators and a clear structure rather than an informal, “free for all” format. One of the participants stated:

> It was curated. It wasn’t just like “a free-for-all.” And that’s really what made it effective. But if there was some way to continue with the program…I don’t know how that would happen on a volunteer basis, you know, with the members of the group.

#### Physical and psychological barriers for implementation require creative solutions

Implementation at the library site presented both physical advantages and challenges for the facilitators. The venue offered suitable space, quiet surroundings, good internet access, and kitchen facilities, but the unexpectedly large group created logistical challenges, including the need to rearrange furniture, manage arrivals, and handle frequent interruptions. Facilitators responded by reorganizing the space to support focus and engagement and by introducing practical solutions such as name tags and structured use of breaks. They also noted the need for clearer rules about mobile phone use and for a gatekeeper to reduce disruptions during sessions. During Session 2, a circular seating arrangement was implemented to ensure focus on the session theme and user engagement.

Participant psychological barriers, which appeared in a couple of sessions, created some worries for the group when one of the participants expressed distressing thoughts, such as the urge “to hit someone in the spine” or “to jump from a bridge.” Facilitators interpreted these statements in light of the speaker’s possible psychological or cognitive problems and managed the session by remaining calm, drawing on clinical expertise, and supporting the group’s accepting response. Hearing such difficulties and recognizing the presence of quieter or less-engaged participants required continual adjustments to communication strategies. Facilitators described feeling uncertainty about how far to encourage participation without applying undue pressure. The question to consider was whether these participants were present because of the incentives, whether they preferred “to listen” over talking and sharing, or whether they had unidentified cognitive or emotional problems. One facilitator reflected:

> I was struggling a bit with how much I could challenge a [person] in the group. On one hand, [they] need to be challenged to move out of passivity; however, when I challenge [them], [they] often offer general answers, and I get a feeling that [they’re] not really sharing how or what [their] plans are. I am afraid of “pushing” too much by asking [them] to be more concrete. For example, I would like to ask [them] what [their] plans are when it comes to returning to work.

These reflections illustrated some of the challenges in the group’s leadership and the need for facilitators to combine professional knowledge from their BKP training with strong communication skills, underscoring the importance of skilled facilitation in future implementations.

#### The online format has advantages and challenges

Transitioning the BKP to an online format during Sessions 4-7 introduced both advantages and challenges. Because facilitators were initially concerned about attendance and technical barriers, participants received detailed instructions, reminders, and targeted follow-up, and an IT assistant supported a hybrid solution at the library. These measures were described as essential to preventing a larger number of dropouts. One facilitator reflected on the transition from face-to-face sessions to online:

> About half of the participants went to the library to get assistance from the IT expert. We started the meeting only a couple of minutes late, which was miraculous. Without the IT assistance, this could have been a huge disruption. I was surprised that the technical problems were overcome so quickly and that we could start the sessions as planned.

Participants generally appreciated the flexibility of attending online, particularly when ill or without transport, and some preferred receiving technical support at the library rather than from family. At the same time, facilitators observed that the first online sessions had less energy and focus than the in-person meetings, and that body awareness exercises were harder to facilitate online until the pace was slowed and more intentional guidance was used.

Participants appeared to be more engaged as well, confirming that the exercise must be slowed down in an online environment.

Facilitators underscored the need for practical adjustments to optimize the hybrid format, such as discouraging simultaneous phone and computer logins, rearranging chairs to improve on-screen visibility, and reconsidering session length to allow sufficient time for reflection and sharing. The 2-hour timeframe seemed insufficient to complete all the learning activities and allow all participants to share. One of the facilitators wrote: “I feel like I rushed through the exercise when I could have offered more prompts to help the participants develop their thoughts and explanations.”

Facilitators also emphasized that reliable technical support and a visible leadership presence—especially in key sessions such as the last meeting—were crucial to maintaining engagement and ensuring a sense of closure across both face-to-face and online formats. The facilitator who lived a 3-hour distance from the library concluded:

> I decided that I had to go to New York to facilitate the last session and ensure the participants were supported and the post-surveys were completed because it was a critical point in the successful completion of the project. The majority of the participants showed up again, and most of them attended in the library.

A couple of participants said they wanted to attend the last meeting in person to close out the experience. The facilitator, who attended the last four sessions from another country, wondered if participants attended the library for the last session because the US-based facilitators had told them that she would be there. Furthermore, she noted that a routine should be established where the first three sessions are face-to-face, the next four are held online, and the seventh and last session is held on-site by one of the facilitators.

### Theme 3: Demand and ideas for continued implementation

Participants reflected on external and internal factors that motivated them to engage and adhere to the intervention. They also came up with ideas on continuation, recruitment, and improvements for future implementation. The theme is further elaborated in three subthemes: (1) The impact of incentives; (2) Expressed need for continuation and ideas for recruitment; and (3) Suggestions for program change, additional content, and methods.

#### Incentives impacted participation

Covering transport costs and providing refreshments at the library helped make participation feasible and created a welcoming atmosphere. Facilitators observed that the little café functioned well, made people feel welcome, and helped participants stay for the whole session. Participants agreed that the financial incentive and gift card initially attracted some people, but they emphasized that most stayed because they experienced the program as genuinely helpful and appreciated the emerging group community. One of the participants reflected:

> A lot of people came because of the incentives and found out it benefited them more than the incentive, but the incentive brought them there, and by the seven weeks we’re still coming. So, it worked out.

Several participants described a shift from “coming for the money” to attending because they valued the support, learning activities, and sense of responsibility to the group. They added that incentives mainly worked as a gateway to meaningful engagement. At the sixth session, one of the participants put it this way: “Before, it was all about the money, but now, it is about the program. I really need this program.”

#### Participants expressed a need for continuation and ideas for recruitment

Upon program completion, participants asked for opportunities for follow-up and further networking within the group but expressed some concerns about how this should be led. Furthermore, the structure of the BKP was highlighted as important, and its possible absence in the follow-up would be a challenge. One suggestion that surfaced was to include some action themes that connected with the follow-up.

> A lot of people have expressed a desire to continue the group, if people wanted to band together and form a support group of some sort. And I don’t know how that would happen, or who would even lead that, because I think the power of the group was that there was a structure, and you guys were the guides and pulled it together, you know. It was curated. Everything was really perfect, but maybe some action items could be good. Maybe just list off one or two or three things that we would hope to take with us or continue, and then some kind of follow-up.

Several participants said they would recommend the BKP to others living with chronic illness and were eager to help “send others to the program.” They proposed broad recruitment efforts targeting, for example, senior communities, outpatient clinics, mental health services, disease-specific organizations, and even people without a diagnosis who are interested in their health. They stressed the importance of presenting the BKP as a mutually shared experience within a caring community, as one participant described: “To let people know that it’s a learning and sharing experience, you know. It does both. It supports, but you also learn. You are supported within a caring community.”

The facilitators reflected on possibly improving the recruitment strategies, including making the assessment of eligible participants more precise and developing a more appropriate question to screen those who might be disruptive to the group, such as “Do you have any emotions or memory issues that would hinder you from showing up to each meeting and communicating with the group?”

#### Suggestions for program change, additional content, and methods were offered

Overall, the participants shared the view that the program should continue as is, with minor adjustments for the US context, such as adding discussions of specific stressors people face in the US and explicit discussions of stress-management strategies. Participants also recommended continuing the BKP in its current format and expressed interest in recommending the program to others with health concerns. Another suggestion was to establish a certificate of completion. Furthermore, they emphasized the need for clear communication and emotional support during the transition from in-person to online sessions, including earlier and more gradual training on the digital platform. One participant put it this way: “You could tell people: You can do it, too, and we’re gonna help you.”

The facilitators identified several areas for refining content and methods. They proposed developing a recorded relaxation exercise to share after introducing it in the group, providing participants with ongoing access at home. In addition, one facilitator noted that the self-selected physical activity was only briefly mentioned in the opening announcements; instead, she should have asked if anyone wanted to discuss their activity progress or challenges, i.e., “I’m available if you want to speak to me about your home activity or speak with me about your activity goals.” The other facilitator, who was responsible for introducing the intervention’s content, recommended streamlining the booklet by reducing or making some questions optional, and revising or removing items that did not work well in practice, to allow more time for meaningful dialogue during sessions.

### Post-intervention survey

Most participants reported that the BKP offered appropriate and useful content, was well organized and appropriately timed, and introduced new approaches to managing their health (Table 5). Ninety-three percent of participants agreed that the program taught them to listen to their body’s signals and use that knowledge to cope and manage. Everyone was satisfied with their ability to talk with the BKP facilitators and would recommend the program to others. Participants strongly agreed on the program’s value and efficacy across the domains of content, materials, organization, communication, and new discoveries.

**Table 5.**
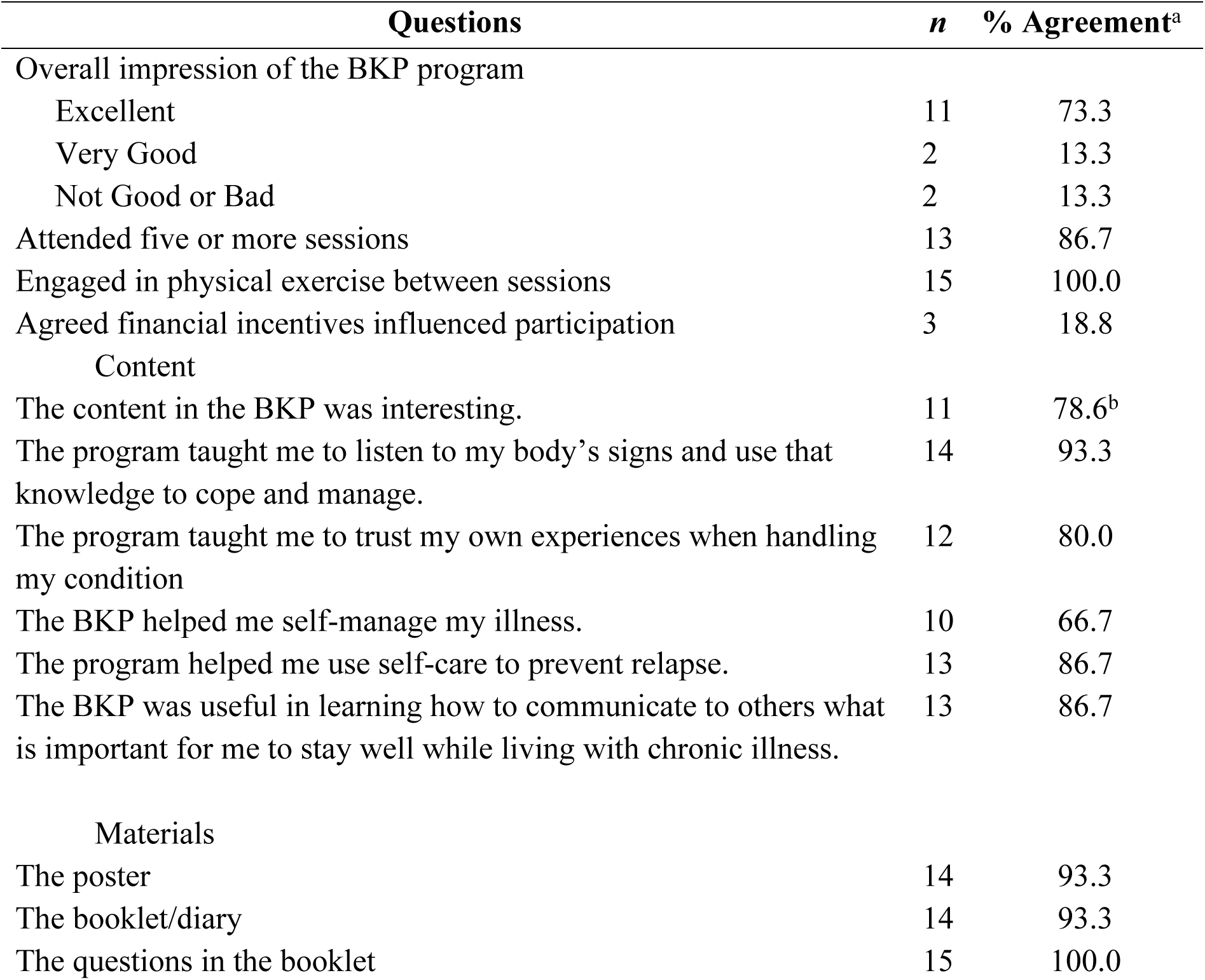
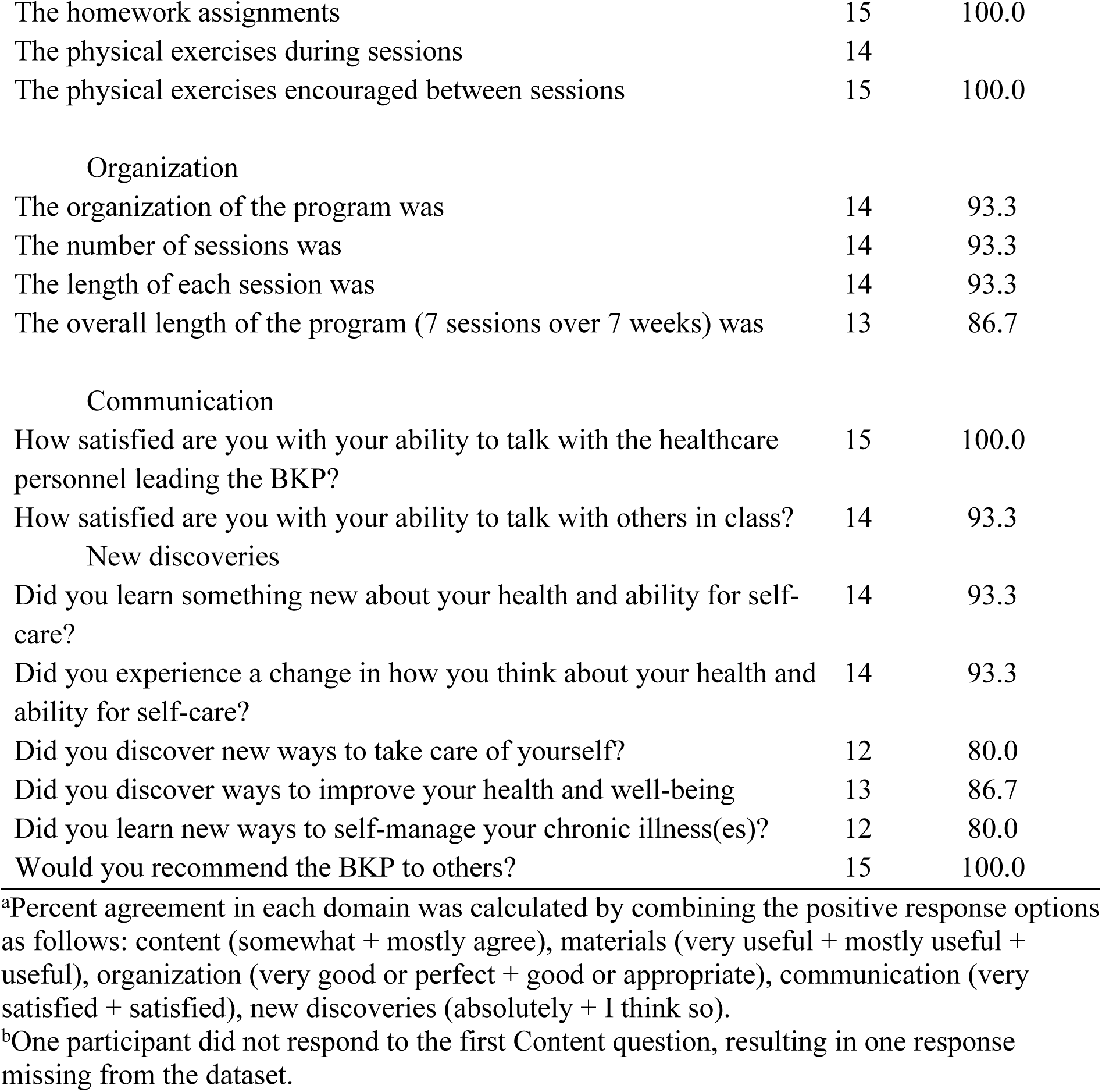
Summary of Post-intervention Survey (n = 15)

| Questions | n | % Agreement<sup>a</sup> |
| --- | --- | --- |
| Overall impression of the BKP program |  |  |
| Excellent | 11 | 73.3 |
| Very Good | 2 | 13.3 |
| Not Good or Bad | 2 | 13.3 |
| Attended five or more sessions | 13 | 86.7 |
| Engaged in physical exercise between sessions | 15 | 100.0 |
| Agreed financial incentives influenced participation | 3 | 18.8 |
| Content |  |  |
| The content in the BKP was interesting. | 11 | 78.6<sup>b</sup> |
| The program taught me to listen to my body's signs and use that knowledge to cope and manage. | 14 | 93.3 |
| The program taught me to trust my own experiences when handling my condition | 12 | 80.0 |
| The BKP helped me self-manage my illness. | 10 | 66.7 |
| The program helped me use self-care to prevent relapse. | 13 | 86.7 |
| The BKP was useful in learning how to communicate to others what is important for me to stay well while living with chronic illness. | 13 | 86.7 |
| Materials |  |  |
| The poster | 14 | 93.3 |
| The booklet/diary | 14 | 93.3 |
| The questions in the booklet | 15 | 100.0 |
| The homework assignments | 15 | 100.0 |
| The physical exercises during sessions | 14 |  |
| The physical exercises encouraged between sessions | 15 | 100.0 |
| Organization |  |  |
| The organization of the program was | 14 | 93.3 |
| The number of sessions was | 14 | 93.3 |
| The length of each session was | 14 | 93.3 |
| The overall length of the program (7 sessions over 7 weeks) was | 13 | 86.7 |
| Communication |  |  |
| How satisfied are you with your ability to talk with the healthcare personnel leading the BKP? | 15 | 100.0 |
| How satisfied are you with your ability to talk with others in class? | 14 | 93.3 |
| New discoveries |  |  |
| Did you learn something new about your health and ability for self-care? | 14 | 93.3 |
| Did you experience a change in how you think about your health and ability for self-care? | 14 | 93.3 |
| Did you discover new ways to take care of yourself? | 12 | 80.0 |
| Did you discover ways to improve your health and well-being | 13 | 86.7 |
| Did you learn new ways to self-manage your chronic illness(es)? | 12 | 80.0 |
| Would you recommend the BKP to others? | 15 | 100.0 |

<sup>a</sup>Percent agreement in each domain was calculated by combining the positive response options as follows: content (somewhat + mostly agree), materials (very useful + mostly useful + useful), organization (very good or perfect + good or appropriate), communication (very satisfied + satisfied), new discoveries (absolutely + I think so).
<sup>b</sup>One participant did not respond to the first Content question, resulting in one response missing from the dataset.

Open-ended questions from the evaluation survey, which yielded 72 comments, also reflected participants’ satisfaction with the materials and exercises, particularly noting that they were informative and thought-provoking and also encouraged self-reflection and awareness of symptoms (S2 Table).

## Discussion

The aim of this study was to assess the feasibility of implementing the Bodyknowledging Program (BKP) at the community level in the US; evaluate participants’ experiences, program components, and self-management outcomes among adults living with chronic illness; and identify the program’s strengths and areas for cultural adaptation to inform its cross-national transferability. Three main themes summarized the findings (Table 4). Overall, the findings of this study were largely consistent with previous research evaluating the BKP intervention and participant outcomes, conducted in Norway. In the following discussion, themes and subthemes identified in this study are compared with previous research on the implementation of the BKP in Norway and other relevant studies.

In Theme 1, US participants clearly conveyed the acceptability of the BKP intervention’s content and approaches and described various outcomes, consistent with the results from Norway. For example, “discovering one’s own health promotion process—acknowledging one’s own body and skill application” aligned with “changing perspectives on health and illness and new ways of thinking and acting towards the illness,” while the subtheme “learning tools are catalysts for self-reflection and empowerment” aligned with two other themes from former studies: “changes in self-awareness, accepting the limits of one’s capacity and regaining control” and “understanding the situations, choices, and actions that make the health condition better or worse” [15, 16].

The participants in the present study highlighted the program’s message of self-empowerment as a central and meaningful part of their experience, particularly how it encouraged them to listen to their bodies, reflect on their health, and develop practical strategies for managing the challenges of living with chronic illness. Similarly, Norwegian participants in previous studies reported that engagement in the BKP improved their ability to manage illness-related problems, leading to greater awareness and utilization of personal resources for health [13]. Furthermore, the US participants conveyed that the BKP supported the development of coping strategies, helped them regain a sense of control, and facilitated their self-management in everyday life, which aligned with Norway’s research outcomes [15, 16]. Both the US and the Norwegian participants reported discovering and utilizing their personal resources for health and becoming more physically and socially active during the intervention period, underscoring the program’s relevance as a health promotion initiative among people living with chronic illness [15, 17, 28]. These findings suggested that the core experience of capacity strengthening for chronic illness self-management was consistent across cultures.

The BKP is based on participants assessing their responses to long-term illness and discovering how they can alter their situation by using their own resources and those in their environment. By contrast, the starting point for the well-known lay-led Chronic Disease Self-Management Program (CDSME) [28] is problem-based and relies heavily on the medical management of a condition. Instead, the BKP presents patient-centered expertise (the phases of Bodyknowledging) for interpretation and application by new patients participating in the program. This approach is in keeping with Paulo Freire’s “pedagogy of the oppressed,” in which the person in question defines their situation and dialogue serves as the main method for helping people to understand their situations and act in new ways. This seems to have a liberating effect because the solutions come from the person and are not based on predefined skills. While the CDSMP [28] targets empowerment through the acquisition of five core self-management skills aimed at behavior change, the BKP approach employs structured learning tools, a reflective booklet, and Bodyknowledging theory visualized in “the rainbow model” [10, 11] (S3 Fig) to support iterative self-assessment, track health processes, and translate bodily signals into new personalized self-management strategies. Across US and Norwegian contexts, participants consistently reported enhanced understanding of their bodily responses, adjusted their activity levels, and reported greater confidence in their own capabilities and broader involvement in meaningful activities [10, 13, 18, 19].

Evident in both the Norwegian and US samples was a gain in greater understanding of factors that made participants’ health conditions better or worse and led them to become more active. Further, peer reflection fostered a sense of belonging and emotional relief [16–19]. These similarities suggested that the central process underlying the BKP may operate similarly across different health service contexts. Such tools may serve as key mediators of empowerment, translating abstract concepts into actionable steps. Nevertheless, the cultural adaptability of these tools warrants further exploration, particularly for diverse populations and varied healthcare settings.

This aligned with Hajdarevic et al.’s [29] narrative review of qualitative studies across chronic illnesses. The researchers note that meaning-making and identity reconstruction in chronic illness are influenced by social norms, relational expectations, and healthcare cultures, while Acuña Mora et al. [30] highlighted potential barriers related to professional roles and system structures within cardiovascular care. Consequently, further research is warranted to explore how the BKP’s reflective tools and visual models can be implemented adaptively across diverse populations and healthcare settings while preserving their core empowerment mechanisms. Participants consistently valued the program’s focus on self-empowerment, describing it as a novel and transformative experience, compared to traditional healthcare encounters based on the biomedical model, which often rely on medication and procedural interventions. There is a gap in current practice, as patients rarely receive structured guidance to leverage their experiential knowledge to support behavior-related changes [8]. The BKP approach addresses this gap by fostering autonomy and self-agency, which are known predictors of improved health outcomes and quality of life [8, 30, 31].

The strong emphasis on peer interaction emerged as a distinctive feature of the BKP, offering participants a sense of belonging and validation that is often absent in conventional care settings. Unlike traditional support groups, the BKP framed discussions around strengths and wholeness rather than illness, fostering resilience and mutual learning. This suggested that group-based interventions can serve as powerful vehicles for empowerment, provided they create safe spaces for sharing and emphasize positive identity reconstruction. Better outcomes from group-format interventions were demonstrated in a systematic review of empowerment interventions in chronic illness [32]. Facilitator strategies, group size considerations, and structured breakout sessions may further enhance interaction and learning in virtual formats. Healthcare professionals who have led BKP groups in Norway emphasize the value of a patient-centered conceptual framework that enables addressing the whole person and individual needs, as well as facilitating patient activation through dialogue-based support [18, 19]. The US participants highlighted the facilitators’ crucial role, presence, and compassion as important components in ensuring a positive outcome. This aligned with studies conducted in Norway [13], underscoring postgraduate training as a prerequisite for leading the BKP. A manual for healthcare professionals, an online training module, and simulation workshops are now established to ensure quality and fidelity in future implementation.

One possible difference between the two contexts was that participants in this study more explicitly compared the program with their usual healthcare experiences. Many described the program’s emphasis on bodily knowledge and self-empowerment as largely absent in their current medical encounters. Such contrasts were less visible in the Norwegian studies, which may reflect differences in healthcare contexts. Another notable difference was that, in the US sample, the majority were women, whereas men and women were equally represented in the Norwegian studies. As the men reported good utility of the BKP in Norway, it is recommended to enroll comparable numbers of male and female representatives in future studies.

Implementation considerations emerging from Theme 2 and the BKP evaluation questionnaire (Table 5 for quantitative and S2 Table for qualitative feedback) demonstrated high participant satisfaction with both program delivery and communication with facilitators. These findings aligned with prior Norwegian studies [15, 18, 19]. A notable adaptation in this US pilot was the condensed engagement structure, consisting of seven consecutive weeks rather than the Norwegian model of seven sessions over 12 weeks [11]. While shorter program durations have been adopted in other chronic disease self-management interventions to reduce participant burden, such formats may also intensify content delivery and increase cognitive and logistical demands [33, 34]. Despite this compressed timeline, participants reported sustained engagement, suggesting that the BKP core components remained intact and transferable.

Another distinguishing feature of the US pilot was the integration of online technology during Sessions 4-7, creating a hybrid delivery model that differed from earlier BKP iterations. The transition to remote participation introduced additional implementation demands, including digital literacy; secure internet access; working electronics (phone, computer, or tablet); accessible email accounts; and online meeting capability. Similar to experiences reported in broader chronic care and digital health interventions, technology-related barriers emerged early during screening and enrollment, prompting the hiring of a dedicated technology support person [35, 36]. Technical considerations were discussed with the participants in the lead-up to the transition to full remote in Session 4. Approximately half of the participants preferred to continue in person, either to maintain community relationships or to obtain access to or competence with technology.

These findings echoed chronic care literature, suggesting that while older adults may be open to using technology to increase convenience and support their activities, successful uptake frequently depends on tailored training, ongoing assistance, and perceived usefulness [35, 37]. In line with this, the hybrid BKP format with on-site technical assistance proved essential in maintaining participation and minimizing dropout. However, this implementation also revealed important logistical challenges. Using a projector to display remote participants caused audio issues, including low volume and feedback, which disrupted communication for both in-person and online groups. Future implementations must thoroughly plan transitions from face-to-face to online delivery, with technological considerations included in the program guide for facilitators. Recruitment efforts will benefit from assessing the technology competencies of people with chronic disease before program onset.

While the overall acceptability of the BKP was high, the findings revealed that older adult participants had mixed experiences with technology. Although some participants viewed digital literacy as a barrier, others emphasized motivation to learn to use new technology as a key determinant for engagement. These insights highlight the need for tailored support and possibly hybrid delivery models to ensure inclusivity. Other studies showed success with hybrid formats and digital health technologies, noting facilitators such as accessibility and motivation, and barriers such as limited digital literacy, limited access, and privacy concerns [35, 36]. This US pilot study demonstrated that the BKP can be delivered in a hybrid format, with positive health impacts and meaningful engagement, through program content, materials, organization, communication, and discoveries which were comparable to those reported in prior research [11, 16].

Financial incentives were used to support recruitment and retention in this multi-week pilot intervention, reflecting variability in healthcare access and utilization in the US and established trends in cultural acceptance and the efficacy of incentive-based approaches to encourage healthy behavior change [38]. Future iterations may reconsider incentives because only a few participants reported that financial incentives influenced their participation. A review of public institutions and insurance companies in upper-middle- and high-income countries revealed that, while incentivized prevention programs demonstrated positive effects on targeted behaviors, there were concerns about sustaining behavior change once incentives were removed [39].

### Strengths and limitations

The BKP was successfully implemented on the community level in a new US cultural context with a socioeconomically and culturally diverse sample. The retention rate was high, and most participants chose to engage in the post-intervention focus group to evaluate the intervention. Some limitations warrant consideration. First, men were not equally represented in the sample; therefore, the relevance for men should be further studied. Second, documenting and reporting chronic illness histories for participants with multiple comorbidities proved challenging. Third, limited computer literacy presented barriers; approximately half the group required assistance during sessions to access email, complete surveys, and navigate technical requirements. Participants also needed support with pre- and post-survey completion, and two participants did not complete the surveys within the allotted 75 minutes. One participant insisted on completing the surveys at home. Future iterations should consider the advantages and disadvantages of moving survey completion to the end of Session 1 and explore options for storing pre-drafted emails to facilitate technical communication during sessions.

## Conclusion

The BKP intervention was well received and acknowledged as a novel and valuable program for strengthening empowerment, health, and well-being among people living with chronic illness in the US, and the acceptability of its health promotion content and approaches was evident in this sample of US participants. The mix of face-to-face and hybrid formats required technical support, which was paramount to enable participants to continue attending online sessions; this should be described as a requirement in the BKP manual. Insights gained relate to improving patient-centered interventions for chronic illness management and informing future large-scale intervention studies.

## Data Availability

Although anonymized, the qualitative transcripts include detailed accounts from a small community sample, creating a risk of indirect re-identification. In addition, participants did not consent to unrestricted public sharing of full transcripts, and such deposition would not comply with ethics committee approval. Relevant supporting materials are available from the corresponding author upon reasonable request.

## Supporting information

**S1 File. Semi-structured Interview Guide on the Feasibility and Acceptability of the Bodyknowledging Program and Process Evaluation.** (DOCX)

**S2 Table. Summary of Evaluation Survey Open-Ended Questions.** (DOCX)

**S3 Fig. Bodyknowledging Theory Visualized in “The Rainbow Model.”** Alt text: Image of “The Rainbow Model” visualization of Bodyknowledging Theory that serves as the theoretical framework and content of the Bodyknowledging Program (BKP) as found in Chapter 16 of Heggdal K. Health promotion among individuals facing chronic illness: the Unique contribution of the Bodyknowledging Program. In Haugan G, Eriksson M, eds., *Health Promotion in Health Care—Vital Theories and Research.* Springer International; 2021: 209–226. (DOCX)

## References

1. European Observatory on Health Systems P. State of Health in the EU Norway: Country Health Profile 2021. OECD Publishing.: 2021.

2. Watson KB, Wiltz JL, Nhim K, Kaufmann RB, Thomas CW, Greenlund KJ. Trends in multiple chronic conditions among US adults, by life stage, behavioral risk factor surveillance system, 2013–2023. Preventing chronic disease. 2025;22:E15. DOI:10.5888/pcd22.240539

3. Larsen PD. Lubkin’s chronic illness: impact and intervention: Jones & Bartlett Learning; 2021.

4. Dehkordi AH, Dehabadi EZ, Rezaei MR, Dehkordi AH, Fattahi F, Oskui AG, et al. Empowerment and self-efficacy in patients with chronic disease; a systematic review study. Journal of Nephropharmacology. 2023;12(2):e10596-e. DOI: 10.34172/npj.2023.10596

5. Organization WH. Patient empowerment and health care In: WHO guidelines on hand hygiene in health care: First global safety challenge clean care is safer care. Geneva World Health Organization, 2009.

6. Vainauskienë V, Vaitkienė R. Enablers of patient knowledge empowerment for self-management of chronic disease: an integrative review. International journal of environmental research and public health. 2021;18(5):2247. DOI 10.3390/ijerph18052247

7. Stepanian N, Larsen MH, Mendelsohn JB, Mariussen KL, Heggdal K. Empowerment interventions designed for persons living with chronic disease - a systematic review and meta-analysis of the components and efficacy of format on patient-reported outcomes. BMC Health Services Research. 2023;23(1):911. doi: 10.1186/s12913-023-09895-6.

8. Varela AJ, Gallamore MJ, Hansen NR, Martin DC. Patient empowerment: a critical evaluation and prescription for a foundational definition. Frontiers in Psychology. 2025;15:1473345. DOI: 10.3389/fpsyg.2024.1473345

9. Van de Velde D, De Zutter F, Satink T, Costa U, Janquart S, Senn D, et al. Delineating the concept of self-management in chronic conditions: a concept analysis. BMJ open. 2019;9(7):e027775. DOI: 10.1136/bmjopen-2018-027775

10. Heggdal K. ’We experienced a lack of tools for strengthening coping and health in encounters with patients with chronic illness’: bridging theory and practice through formative research. International Practice Development Journal. 2015;5(2). DOI: 10.19043/ipdj.52.004

11. Heggdal K. Health promotion among individuals facing chronic illness: The unique contribution of the bodyknowledging program. Health promotion in health care–vital theories and research. 2021:209–26. DOI: 10.1007/978-3-030-63135-2_16

12. Heggdal K. Kroppskunnskaping: en grunnleggende prosess for mestring av kronisk sykdom: [Bodyknowledging: A Basic Process for Coping with Chronic Illness] [PhD thesis]. Faculty of Medicine, Institute of Social Medicine, Department of Nursing Science. Norway: University of Bergen; 2003.

13. Heggdal K, Lovaas BJ. Health promotion in specialist and community care: how a broadly applicable health promotion intervention influences patient’s sense of coherence. Scandinavian Journal of Caring Sciences. 2018;32(2):690–7. DOI: 10.1111/scs.12498

14. Heggdal K. Utilizing bodily knowledge in patients with chronic illness in the promotion of their health: a grounded theory study. California Journal of Health Promotion. 2013;11(3):62–73. DOI: 10.32398/cjhp.v11i3.1542

15. Heggdal K. Patient’s experience of the outcomes of engaging in a broadly applicable health promotion intervention for individuals facing chronic illness. Health. 2015;7(6):765–75. DOI: 10.4236/health.2015.76091

16. Engevold MH, Heggdal K. Patients’ experiences with changes in perceived control in chronic illness: a pilot study of the outcomes of a new health promotion program in community health care. Scandinavian Psychologist. 2016;3. https://psykologisk.no/sp/2016/03/e5/

17. Heggdal K, Oftedal B, Hofoss D. The effect of a person-centred and strength-based health intervention on recovery among people with chronic illness. European Journal for Person Centered Healthcare. 2018;6(2):279–85. DOI: 10.5750/ejpch.v6i2.1461

18. Heggdal K, Mendelsohn JB, Stepanian N, Oftedal BF, Larsen MH. Health-care professionals’ assessment of a person-centred intervention to empower self-management and health across chronic illness: Qualitative findings from a process evaluation study. Health Expectations. 2021;24(4):1367–77. DOI: 10.1111/hex.13271

19. Heggdal K, Stepanian N, Oftedal BF, Mendelsohn JB, Larsen MH. Health care professionals’ experiences of facilitating patient activation and empowerment in chronic illness using a person-centered and strengths-based self-management program. Chronic illness. 2023;19(1):250–64. DOI: 10.1177/17423953211065006

20. Gray JR, Grove SK. Burns and grove’s the practice of nursing research-E-book: Appraisal, synthesis, and generation of evidence: Elsevier Health Sciences; 2020.

21. Creswell JW, Poth CN. Qualitative inquiry and research design: Choosing among five approaches: Sage publications; 2016.

22. Antonovsky A. Unraveling the mystery of health: How people manage stress and stay well: Jossey-bass; 1987.

23. McCormack B, McCance T. Person-centred practice in nursing and health care: theory and practice: John Wiley & Sons; 2016.

24. Clarke V, Braun V. Thematic analysis: a practical guide. Thematic analysis. 2021;9:1–100.

25. Kraiwanit T, Limna P, Siripipatthanakul S. NVivo for social sciences and management studies: A systematic review. Advance Knowledge for Executives. 2023;2(3):1–11.

26. Bowen DJ, Kreuter M, Spring B, Cofta-Woerpel L, Linnan L, Weiner D, et al. How we design feasibility studies. American journal of preventive medicine. 2009;36(5):452–7. DOI: 10.1016/j.amepre.2009.02.002

27. Korstjens I, Moser A. Series: Practical guidance to qualitative research. Part 4: Trustworthiness and publishing. European Journal of General Practice. 2018;24(1):120–4. DOI: 10.1080/13814788.2017.13755092

28. Lorig KR, Holman HR. Self-management education: history, definition, outcomes, and mechanisms. Annals of behavioral medicine. 2003;26(1):1–7. DOI: 10.1207/S15324796ABM2601_01

29. Hajdarevic S, Norberg A, Lundman B, Hörnsten Å. Becoming whole again—Caring for the self in chronic illness—A narrative review of qualitative empirical studies. Journal of clinical nursing. 2025;34(3):754–71. DOI: 10.1111/jocn.17332

30. Acuña Mora M, Bratt E-L, Saarijärvi M. Taking charge of your health: enabling patient empowerment in cardiovascular care. European Journal of Cardiovascular Nursing. 2024;23(7):814–7. DOI: 10.1093/eurjcn/zvae015

31. Mortimer JT, Staff J. Agency and subjective health from early adulthood to mid-life: Evidence from the prospective youth development study. Discover social science and health. 2022;2(1):2. DOI: 10.1007/s44155-022-00006-0

32. Costa IG. Engaging older persons in self-management of illness. Supporting Older Persons on Their Aging Journey: An Emancipatory Approach to Nursing Care. 2024;203.

33. Huang Y, Li S, Lu X, Chen W, Zhang Y, editors. The effect of self-management on patients with chronic diseases: a systematic review and meta-analysis. Healthcare 2024;12(21):2151. DOI: 10.3390/healthcare12212151

34. Alliston P, Jovkovic M, Khalid S, Fitzpatrick-Lewis D, Ali MU, Sherifali D. The effects of diabetes self-management programs on clinical and patient reported outcomes in older adults: a systematic review and meta-analysis. Frontiers in Clinical Diabetes and Healthcare. 2024;5:1348104. DOI: 10.3389/fcdhc.2024.1348104

35. Leung DKY, Wong NHL, Yau JHY, Wong FHC, Liu T, Kwok W-W, et al. Hybrid-delivered community psychoeducation for people aged 50 and older: A mixed-method evaluation and lesson learned. Internet Interventions. 2024;35:100699. DOI: 10.1016/j.invent.2023.100699

36. Hepburn J, Williams L, McCann L. Barriers to and facilitators of digital health technology adoption among older adults with chronic diseases: updated systematic review. JMIR aging. 2025;8:e80000. DOI: 10.2196/80000

37. Zhao J, Chen D, Barbareschi G, Sato C, editors. The Role of ICT Tools through a Community Program Co-creation in a Japanese Aging Community. Proceedings of the 2025 CHI Conference on Human Factors in Computing Systems; 2025. 1–10. DOI: 10.1145/3706598.3714158

38. Higgins ST, Silverman K, Sigmon SC, Naito NA. Incentives and health: an introduction. Elsevier; 2012. p. S2–S6. DOI: 10.1016/j.ypmed.2012.04.00

39. Petracca F, Ardito V, Sala F, Bertolaso G, Menichelli L, Vecchio R, et al. Harnessing financial incentives for health promotion: A scoping review of prevention programs implemented in upper-middle and high-income countries. Social Science & Medicine. 2025:118499. DOI: 10.1016/j.socscimed.2025.118499

